# Effectiveness of incentivized peer referral to increase enrollment in a community-based chlamydia screening and treatment study among young Black men

**DOI:** 10.1101/2022.09.09.22279725

**Authors:** Mary Beth Campbell, Aneeka Ratnayake, Gérard Gomes, Charles Stoecker, Patricia Kissinger

**Affiliations:** Tulane University School of Public Health and Tropical Medicine, Epidemiology Department; 1440 Canal St., Suite 2000, New Orleans, LA 70112; Tulane University School of Public Health and Tropical Medicine, Health Policy and Management Department; 1440 Canal St., Suite 2000, New Orleans, LA 70112

**Keywords:** Recruitment, incentivized peer referral, Black men, multiple time series analysis, COVID-19, STI, implementation science, *Chlamydia trachomatis*, Interrupted Time Series Analysis

## Abstract

**Purpose:** Incentivized peer referral (IPR) has been shown to be an effective method of recruitment for men who have sex with men but has not been studied extensively in men who have sex with women (MSW), particularly among Black MSW. We aimed to determine if IPR was more effective than uncompensated peer referral for recruiting young Black men into a community STI screening study.

**Methods:** We used data from the *Check It* study, a chlamydia (Ct) screening and treatment program for young Black men ages 15-26 in New Orleans, LA. Enrollment was compared before and after IPR was implemented using Multiple Series Analysis (MTSA). IPR was introduced to increase recruitment that had been severely diminished because of the COVID-19 shutdown.

**Results:** Of 1527 men enrolled, 1399 (91.6%) were enrolled pre-IPR and 128 (8.4%) were enrolled post-IPR. The percentage of men referred by a friend or peer was higher in the post-IPR period than in the pre-IPR period (45.7% vs. 19.7%, *p*<0.001). Post-pandemic, we observed a statistically significant increase of 2.007 more recruitments (p=0.044, 95% CI (0.0515, 3.964)) at the start of the post-IPR era, compared to the pre-IPR era. Overall, we also observed a trending increase in recruitments in the IPR era relative to the pre-IPR era (0.0174 recruitments/week, *p*=0.285, 95% CI (−0.0146, 0.0493)) with less recruitment decay in the post-IPR compared to pre-IPR.

**Conclusions:** IPR may be an effective means of engaging young Black men in community based STI research and prevention programs, particularly when clinic access is limited.

## Introduction

*Chlamydia trachomatis* (Ct) is the most common reportable sexually transmitted infection (STI) in the United States, with over 1.8 million cases reported in 2019[1]. The state of Louisiana currently ranks 3^rd^ in Ct case rates, with a rate of 775.3 cases per 100,000 population, 40% more than the national rate[2]. Within Louisiana, the greater New Orleans area has the third highest Ct diagnosis rate of all major metropolitan areas in the US[1].

Youth ages 15-24 years old experience the highest burden of Ct infection, accounting for almost two-thirds (61.0%) of all reported US cases in 2019[1]. Black youth have the highest risk of Ct infection[3], and young Black men ages 15-19 have a case rate 9.1 times higher than that of their White counterparts[4]. Young men in this age group, particularly young Black men, are also less likely than their female counterparts to attend regular health care visits and are less likely to be insured[5]. This is of concern as most Ct infections are asymptomatic – thus, screening in non-clinic settings is important in Ct detection and control efforts. Current Ct screening guidelines are focused on young women ages 24 and younger, due to the serious sequelae that can occur from untreated Ct infections, including infertility, pelvic inflammatory disease, and ectopic pregnancy[6-8].

At present, CDC guidelines do not recommend routine screening for young men who have sex with women, stating feasibility of testing men as a reason[9]. Because of this, efforts have been made to determine if there are benefits to increasing screening and treatment in young men. The *Check It* study is a novel, bundled, community-based seek, test, and treat Ct screening program for 15-to 26-year-old Black men in New Orleans who have sex with women (MSW)[11]. *Check It* was designed with the purpose of determining if community-based Ct screening of young Black men ages 15-26 in New Orleans could both address barriers to Ct screening and treatment in this group and decrease rates of Ct in young women in New Orleans.

Engaging young Black men in medical services and research has long been a challenge, largely due to historical trauma and experienced racism in the US medical system[12]. Young Black men disproportionately experience greater barriers to healthcare. These barriers exist at multiple levels, including structural (e.g., transportation, inconvenient clinic hours, costs), individual (e.g., competing priorities, lack of time, confidentiality/privacy concerns, stigma, fear, low-risk perception), cultural (e.g., feelings of invincibility, masculinity norms, reluctance to seek health care), discrimination (e.g., individual, structural), and interpersonal (e.g., upbringing, negative influencers)[13-16]. To help mitigate any barriers to study recruitment, study participants were encouraged to refer their peers to the study as part of the study protocol as a means of establishing trust and community. Peer referral became especially important during the COVID-19 pandemic, as the usual barriers to recruitment were exacerbated by a two-month shut-down and continued restrictions on in-person recruitment events.

Incentivized peer referral (IPR) is a form of peer referral in which study participants are compensated for each peer they successfully recruit into the study and has been shown to be an effective means of recruitment for HIV studies and care among men who have sex with men[17-20], though less is known about the effect on recruitment among men who have sex with women, particularly Black young men. Based on this evidence, the *Check It* protocol was revised to include a modest monetary incentive of $5 per peer referral. This IPR approach was implemented in July 2020 to help mitigate the effects of the COVID-19 pandemic on in-person recruitment and was continued through the end of the study. The purpose of this analysis is to determine if this addition of IPR was associated with an increase in peer recruitment and, therefore, if it is an effective means of engaging young Black men to participate in a community-based Ct screening and treatment program.

## Methods

### Population and setting

IPR data was collected via the *Check It* study, a community-based Ct and gonorrhea (GC) screening program in New Orleans for young Black men between the ages of 15 and 26 who have sex with women. The *Check It* study staff recruited participants via both venue-based and social marketing strategies between May 2017 and May 2021. To be eligible for the study, participants had to meet the following eligibility criteria: be assigned male sex at birth (based on the question “do you have a penis?”); identify as African American or Black; be between the ages of 15–26 at enrollment; live or spend most of their time in Orleans Parish; report having had vaginal sex with a woman in their lifetime; be willing and able to consent to study activities; speak and understand English; did not take azithromycin in the 2 weeks prior to enrollment; and did not previously enroll in the *Check It* study. Participants were provided with $25 compensation, in the form of a gift card or service of equivalent value (e.g., haircut, event ticket).

Peer referral data was collected between March 7, 2018 and May 28, 2021. Due to the COVID-19 pandemic citywide shut down, venue-based recruitment was halted March 23, 2020 temporarily.,. Limited field recruitment was reinitiated on May 16, 2020. Though venue-based recruitment resumed after New Orleans COVID stay-at-home orders were lifted, the remaining COVID restrictions and realities of the pandemic continued to render recruitment in the community a challenge. Using evidence of the effectiveness of IPR in other studies, the *Check It* protocol was revised to include IPR and was approved by Tulane University IRB on July 27, 2020.

IPR was conducted between July 28, 2020 and the final day of recruitment, May 28, 2021. Participants were offered $5 for each person they referred and who was successfully enrolled in the study. To further encourage participants to refer friends into the study, this was marketed as “Refer 5 friends for $25 incentive” at recruitment events and on the *Check It* Instagram profile. New participants were introduced to IPR during study consent and enrollment and were asked if a friend or peer referred them to the study. Previously enrolled participants who agreed to future contact were sent an IRB-approved communication via SMS or Instagram direct message describing the IPR program (Figure 1). *Check It* study staff provided strategies for engaging peers that included communication implements such as text messages, printed materials, and sharing *Check It* study information on social media.

**Figure 1:**
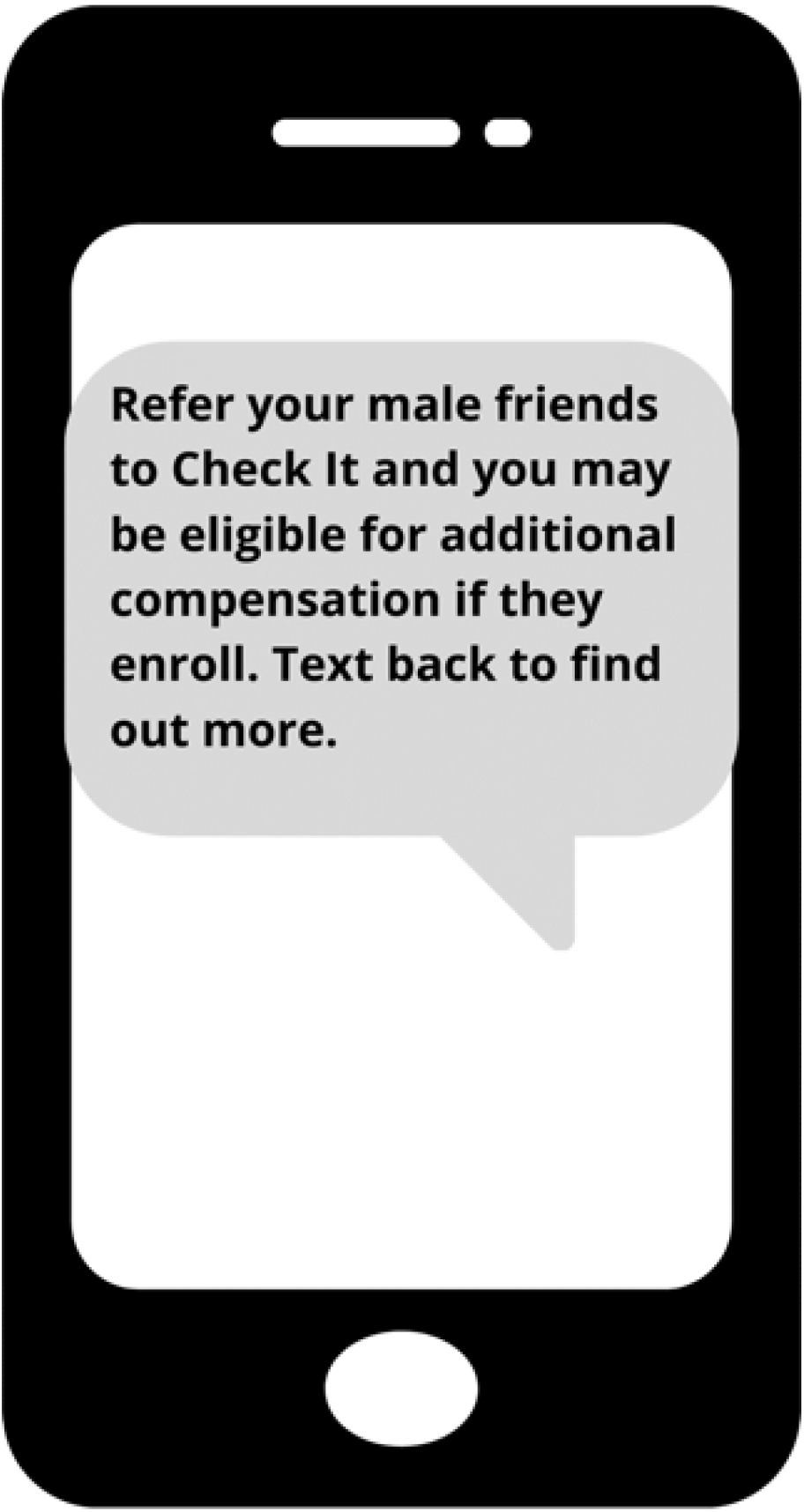
IRB-approved IPR Message.

### Statistical Analysis

Data were limited to that collected between March 7, 2018, the day that collection of peer referral data began, and May 28, 2021, the final day of enrollment. Descriptive statistics were performed for this subset of participants (Table 1) using IBM SPSS Statistics 24 (IBM Corp, Armonk, NY, 2016).

**Table 1:** Participant Characteristics, N = 1534.

| **Table 1: Participant Characteristics, N = 1534** |  |
| --- | --- |
| Characteristics | N (%) |
| Age, years (mean ± SD) | 20.1 ± 2.6 |
| Education level |  |
| Less than high school | 37 (2.4) |
| High school student | 282 (18.5) |
| High school graduate | 570 (37.4) |
| More than high school | 627 (41.1) |
| Other/Refuse to answer | 10 (0.7) |
| Ct+ in parent study |  |
| Yes | 170 (11.1) |
| No | 1343 (87.7) |
| Specimen error | 18 (1.2) |
| Multiple female partners in past 2 months |  |
| Only one female partner | 1271 (63.7) |
| >1 female partner | 724 (36.3) |
| Living situation |  |
| Living at home with at least one parent/guardian | 837 (54.9) |
| Living at home with a foster family | 15 (1.0) |
| Living at a friends' home with at least one parent/guardian present | 36 (2.4) |
| Living on your own (alone or with roommates) | 243 (15.9) |
| Living in a dorm | 205 (13.4) |
| Living at home with parent, but attending college or working | 137 (9.0) |
| No regular place to live | 18 (1.2) |
| Other | 22 (1.4) |
| Refuse to answer | 12 (0.8) |
| Heard about study from a friend/peer |  |
| Yes | 335 (21.9) |
| No | 1192 (78.1) |

The aim of this analysis was to determine if the rate of peer recruitment in the *Check It* study increased after the implementation of IPR, compared to the rate of peer recruitment during the pre-IPR period of the study. IPR was implemented in the final 10 months of the study as a means of increasing enrollment during the COVID-19 pandemic. Prior to the implementation of IPR, peer referral was uncompensated, and between May and July 2020 recruitment strategies overall were adjusted to meet the requirements of the COVID-19 pandemic. Thus, there were three distinct time periods in this study: March 2018-March 2020, pre-pandemic and pre-IPR; May-July 2020, post-pandemic and pre-IPR; and July 2020-May 2021, post-pandemic and post-IPR. Note that the period of March-May 2020 has been omitted, as no recruitments occurred during this time due to COVID-19 lock downs in New Orleans.

As there are pre- and post-intervention periods with multiple observations on the outcome variable recorded during both periods, a multiple time series analysis (MTSA), a variation of the interrupted time series analysis, was conducted. The interrupted time series analysis (ITSA) has increasingly been used to evaluate public health interventions such as IPR[21]. ITSA and MTSA are particularly suited to the evaluation of interventions introduced to a population over a clearly defined period[21]. Using the time series of an outcome of interest to determine an underlying trend, the introduction of an intervention at a specific time or multiple time points -which interrupts this trend-is evaluated to determine its impact on the outcome of interest[21 22]. Thus, this analysis is particularly suited to evaluate the IPR intervention. To account for the observed variability in enrollment, a 7-day rolling average was used, which allowed us to account for daily variation in recruitment numbers (e.g. weekdays vs weekends). In addition, the period from March 20-May 16, 2020 was omitted as no recruitments occurred during this time due to COVID-19 shut-down measures. The ITS analysis was performed using Stata 15.0 (StataCorp, College Station, TX, 2021), utilizing the ITSA code created by Ariel Linden of Linden Consulting Group, LLC[22].

## Results

Between March 7, 2018 and May 28, 2021, 1,534 participants were enrolled into the *Check It* study. Of these participants, 335 (21.9%) had been referred to the study by a friend or peer. Participant characteristics are summarized in Table 1. The mean age was 20.1 years (S.D. 2.6). Participants’ mean number of female sex partners was 14.98 (S.D. 24.25) and 170 (11.1%) of participants tested positive for Ct in the *Check It* parent study. Thirty-seven participants reported having a less than high school education; 282 (18.5%) were current high school students; 570 (37.4%) had graduated high school; 627 (41.1%) reported having more than a high school education; and 10 (0.7%) reported having an education of “Other” or refused to answer. Regarding living situation, most participants (837, 54.9%) reported living at home with at least one parent or guardian. For the remaining participants, 15 (1.0%) lived with a foster family; 36 (2.4%) lived at a friends’ home with at least one parent or guardian present; 243 (15.9%) lived on their own, either alone or with a roommate; 205 (13.4%) reported living in a dormitory; 137 (9.0%) lived at home with a parent but attended college or were working; 18 (1.2%) reported not having a regular place to live; 22 (1.4%) reported their living situation as “Other”; and 12 (0.8%) refused to answer the question.

Participants (n=1391) were enrolled between March 7, 2018 and March 20, 2020, at which time COVID-19 shut-down measures were enacted in New Orleans. In the post-shut-down period between May 17 and July 27, 2020, 13 participants enrolled in the study; it should be noted that peer recruitment was still uncompensated during this period and that only 2 of these participants (15.4%) were referred by a peer. Incentivized peer referral (IPR) was approved by Tulane IRB on July 27, 2020 and implemented on July 28, 2020. The remainder of the participants (129) were enrolled between July 28, 2020 and May 28, 2021 in the post-IPR period. Of these 129 participants, 59 (45.7%) were referred by a peer. As peer recruitment was not only affected by the introduction of the IPR intervention but also the COVID-19 pandemic, multiple group ITSA was conducted to compare recruitment across these three distinct periods. It should be noted that, since no participants were enrolled during the lockdown period of March 20-Mary 16, 2020, this period was omitted from the analysis.

A multiple time series analysis (MTSA) was conducted to examine the impact of IPR on the rate of participant enrollment. MTSA modeling indicates an initial mean of 15.21 (p<0.01, 95% CI (11.176, 19.247)) recruitments per week with a decline of 0.00567 (p=0.222, 95% CI (−0.0148, -0.00347)) enrollments per week over the period before the COVID-19 stay at home orders were put in place. In the post-stay at home/pre-IPR period of May 17-July 27, 2020, there was a sharp drop of -9.091 (p<0.001, 95% CI (−12.855, -5.326)) recruitments in the first week relative to the pre-pandemic period. Over this segment of the pre-IPR period there was a further weekly decline of -0.00992 recruitments per week (p=0.547, 95% CI (−0.0424, 0.0225)). In the post-IPR period of July 27, 2020 until the study’s end on May 28, 2021, there was an increase in the first week of 2.007 recruitments (p=0.044, 95% CI (0.0515, 3.964)) per week followed by a slightly upward, though not significant trend of 0.0174 participants recruited per week (p=0.285, 95% CI (−0.0146, 0.0493)). Enrollment trends, collapsed to weekly totals for visual clarity, are shown in Figure 2.

**Figure 2:**
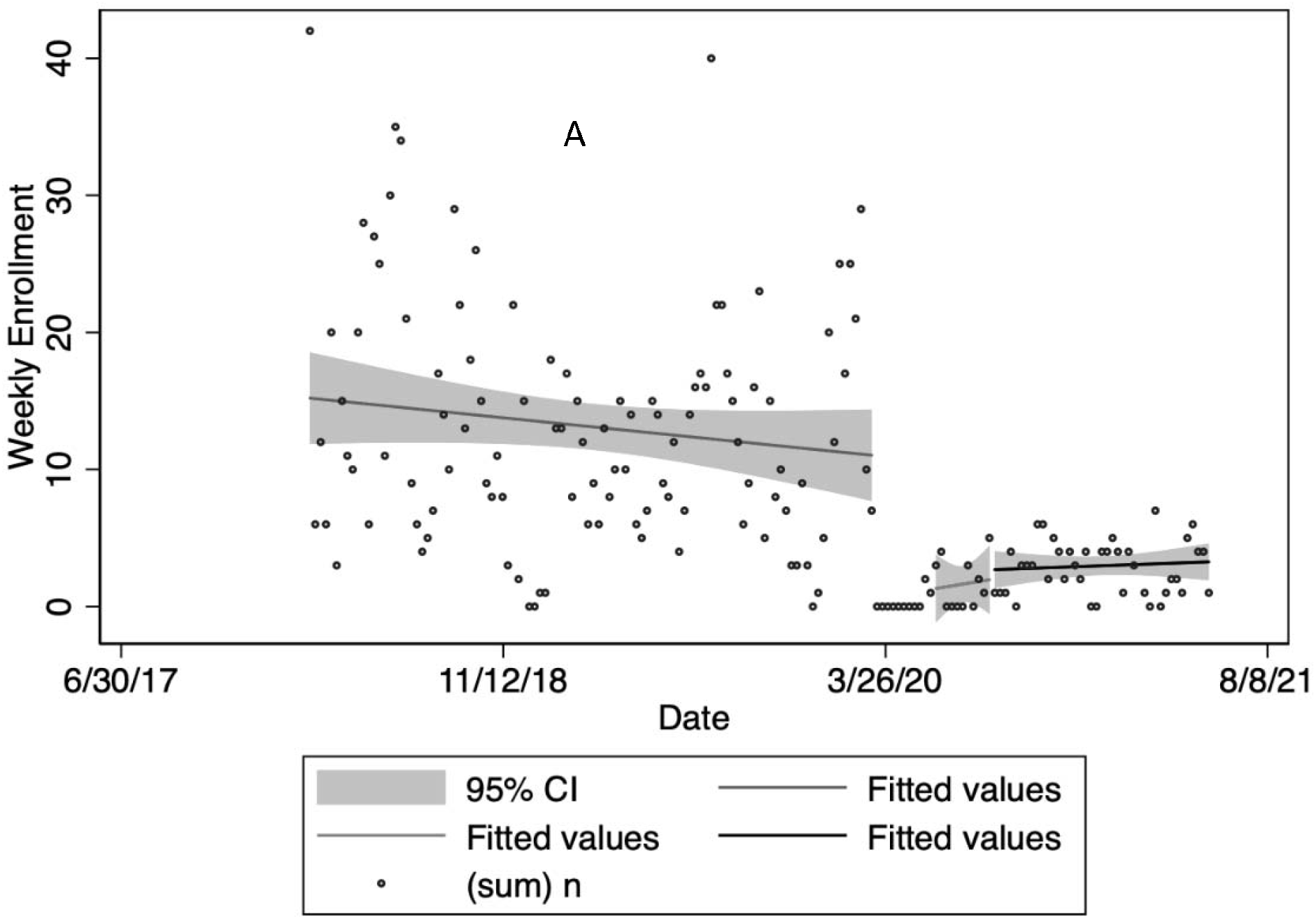
Multiple Times Series Analysis of Recruitment Data Through Transition to IPR. (A) March 2018-March 2020, pre-pandemic and pre-IPR (B) Study temporarily suspended due to COVID-19 (C) May-July 2020, post-pandemic and pre-IPR (D) July 2020-May 2021, post-pandemic and post-IPR

## Discussion

The MTSA indicates that IPR had a slight positive impact on recruitment of young Black men who have sex with women into a chlamydia (Ct) and gonorrhea (GC) seek, test, and treat community program in New Orleans, LA and may have served as a safety net for recruitment during clinic interruptions such as the COVID-19 shutdown. After declines seen in the pre-COVID-/pre-IPR and post-COVID/pre-IPR periods, the implementation of IPR resulted in a small increase in participant recruitment rate (an initial jump of 2.007 recruitments and increasing at a rate of 0.0174 participants recruited per week). While these increases are modest, so too was the cost of IPR at just $5 per recruit. This positive trend is similar to those observed in other community-based studies which used IPR as a means of recruitment, even though those other studies were predominantly focused on HIV, not other STIs, and targeted young men who have sex with men (MSM) and transgender women[18-20].

It must be noted that the rate of participant recruitment was already declining before the COVID-19 pandemic, due in part to saturation at our community sites. IPR could serve to combat decay in recruitment as it serves to increase the potential subject pool, whereas venue-based recruitment is subjected to finite numbers of person frequenting venues.

And, as expected, the results observed in the analysis were further influenced by the COVID-19 pandemic. In the period between the start of stay-at-home orders in March 2020 and approval for IPR, recruitment in general was low. This was because the *Check It* study could no longer hold in-person recruitment events or readily recruit in the community; instead, individuals who were interested in participating had to make an appointment with a study recruiter, which meant that recruitment slowed significantly during this time. Though stay-at-home orders were lifted in May 2020 the recruitment team was still limited in recruitment activities, since many of our community partners remained closed or severely limited in service. No participants were recruited in the stay-at-home period of March 20-May 16, 2020, and in the post-stay-at-home/pre-IPR period of May 17-July 27, 2020 only 13 participant were enrolled. In the post-IPR period of July 28, 2020 through the end of the study on May 28, 2021, only 129 additional participants were enrolled. Though this was an improvement compared to the time between March 20 and July 27, 2020, these numbers and the results of the analysis suggest that a larger sample size is needed to better evaluate the impact that IPR had on recruitment. Moreover a longer IPR implementation period would help us to determine if the decay seen in the pre-IPR would be seen in the IPR as well.

There are a few COVID-19-related factors that should be mentioned to help interpret the data. STI testing services in New Orleans were limited during and after stay-at-home orders were in place, which may have incentivized participants to enroll in the study, particularly those who had symptoms. The loss of job opportunities in New Orleans during the pandemic may have made participants more likely to enroll and refer peers in order to collect the incentive money. Though the results observed in the post-IPR period may have been impacted by the pandemic, it is important to acknowledge that the future of study recruitment is now inextricably linked to both COVID-19 and its consequences. Thus, as we emerge from the pandemic, the impact of IPR on study recruitment is worth studying further.

These results suggest that IPR may be a useful strategy to employ, alongside other efforts, when trying to recruit and engage young Black men in STI research and care. This may be particularly useful in contexts in which in-person recruitment may be limited, including school closures, holidays, and in remote settings with limited access to STI testing and treatment centers. Employing IPR as a recruitment and engagement strategy has the potential to have great clinical and public health implications and could also indirectly contribute to a decrease in STI transmission to their female sexual partners.[23]

Among youth in particular, services referred by peers, especially remote and virtual services, may have become more appealing than in-person clinics. In addition, an increase in virtual opportunities for making money which arose due to the COVID-19 pandemic further justifies the benefits of IPR. Additionally, IPR has the potential to help increase engagement of young Black men in research.

Mistrust of research and medical institutions and investigators has been noted as the most significant barrier to research participation by the Black community[12]. One of the consequences of institutions failing to meet the needs of and engage young Black men in research is that they are underrepresented, and other groups are overrepresented in studies. This underrepresentation, in turn, contributes to increased inequities and health disparities. Institutions and investigators must make many changes in research study design and implementation to mitigate this issue. Peer referral, specifically IPR, is one strategy to take, as evidenced by this study. Individuals who are enrolled in a study that they trust may be more likely to refer their peers, especially if they receive an incentive for doing so. In turn, individuals may be more likely to enroll in a study if they are referred by someone they trust. The literature and the results of this analysis suggest this. It is thus worth studying and implementing IPR in future research studies and outreach programs aimed at young Black men and other underserved groups, especially when the COVID-19 pandemic subsides.

## Supporting information

Equator_checklist

## Data Availability

All data produced in the present study are available upon reasonable request to the authors

## Acknowledgements

This work was funded by the US National Institutes of Health National Institute of Child Health and Human Development/National Institute of Allergy and Infectious Diseases [R01HD086794]. This work was originally presented as an abstract at the 2021 International Society for Sexually Transmitted Diseases Research (ISSTDR) meeting.

